# From migration to mealtime: how income, education, and age of arrival shape Black immigrant mothers’ food parenting practices and their children’s dietary habits

**DOI:** 10.1101/2025.08.27.25334562

**Authors:** Phoebe P. Tchoua, Deborah Quesenberry

## Abstract

Food parenting practices (i.e., feeding practices) play a crucial role in shaping children’s nutritional environment and dietary behaviors, yet little is known about these practices among Black immigrant parents in the U.S. This study examined the food parenting practices of Black immigrant mothers using a modified Comprehensive Home Environment Survey and refined a conceptual model: *Influences of Food Parenting Practices on BMI*. Mothers were recruited using purposive and snowball sampling strategies. We conducted univariate descriptive statistics, independent t-tests, and multiple linear regression analyses. Among the 103 Black immigrant mothers analyzed, most were aged 25–34 years (60.2%), identified as non-Hispanic Black (93.2%), were married (82.5%), had household incomes ≥$50,000 (53.4%), and had migrated to the U.S. at an average age of 14.8±10.1 years from 32 different countries. Across all food parenting practice constructs and subconstructs assessed, there was greater use of pressure to eat (0.65±0.27), meal and snack routines (0.67±0.22), healthy food availability (0.74±0.22), and child autonomy (0.59±0.22). Income, age of migration, education, and the child’s biological sex significantly contributed to models of specific food parenting practices. Higher household income and education levels were associated with greater availability of healthy foods. Participants who migrated after age 14 were more likely to model unhealthy eating behaviors. Female children were more likely to participate in meal planning compared to male children. These findings helped inform the final model, *Influences of Food Parenting Practices on BMI*, with most hypothesized relationships supported and several new pathways emerging. Public health Researchers and practitioners should consider these maternal factors and the developed model when designing nutritional interventions and education targeting this population.

## Introduction

Excess body weight has become increasingly prevalent over the past 30 years, with increased prevalence among Black and African-American (Black) populations.^1,2^ Health conditions and metabolic aberrations related to excess adipose tissue have also become more prevalent.^3–5^ Over a span of 30 years, 1988 and 2018, an overabundance of food, alongside other factors, contributed to an 8.9% increase in obesity prevalence among children and adolescents aged 2-19 years in the United States (U.S.), 10.0% to 18.9%.^6^ Among racial groups during this same time period, Black children experienced an approximate two-fold increase (12.5% to 23.1%) compared to White children (9.2% to 15.1%).^6^ In addition to excessive food intake, a major contributor to obesity, studies have also shown that food parenting practices can influence a child’s BMI and risk of becoming overweight or obese.^7–9^ Food parenting practices are feeding-specific styles and practices that parents use with their children and consist of three major constructs, coercive control, structure, and autonomy support or promotion.^10^

The Social Cognitive Theory (SCT) provides a framework for understanding health behaviors related to nutrition^11^ and serves as the foundation for this research. SCT posits that behavior is shaped by triadic reciprocal factors: cognitive/personal, behavioral, and environmental. In many families, mothers are primarily responsible for feeding their children and therefore influence their children’s BMI in several ways. As shown in the proposed theoretical model, *Influences of Food Parenting Practices on BMI* (see Fig 1), a mother’s knowledge, attitudes, and beliefs, along with her acculturation toward nutrition influence her food parenting practices. Within this framework, food parenting practices function as behavioral factors that directly influence a child’s dietary habits. Focus group findings with Black immigrant mothers confirm that a mother’s knowledge, attitudes, and beliefs (i.e., cognitive/personal), along with her level of acculturation (i.e., environmental), shape these practices (see Fig 1).^12^

**Fig 1.**
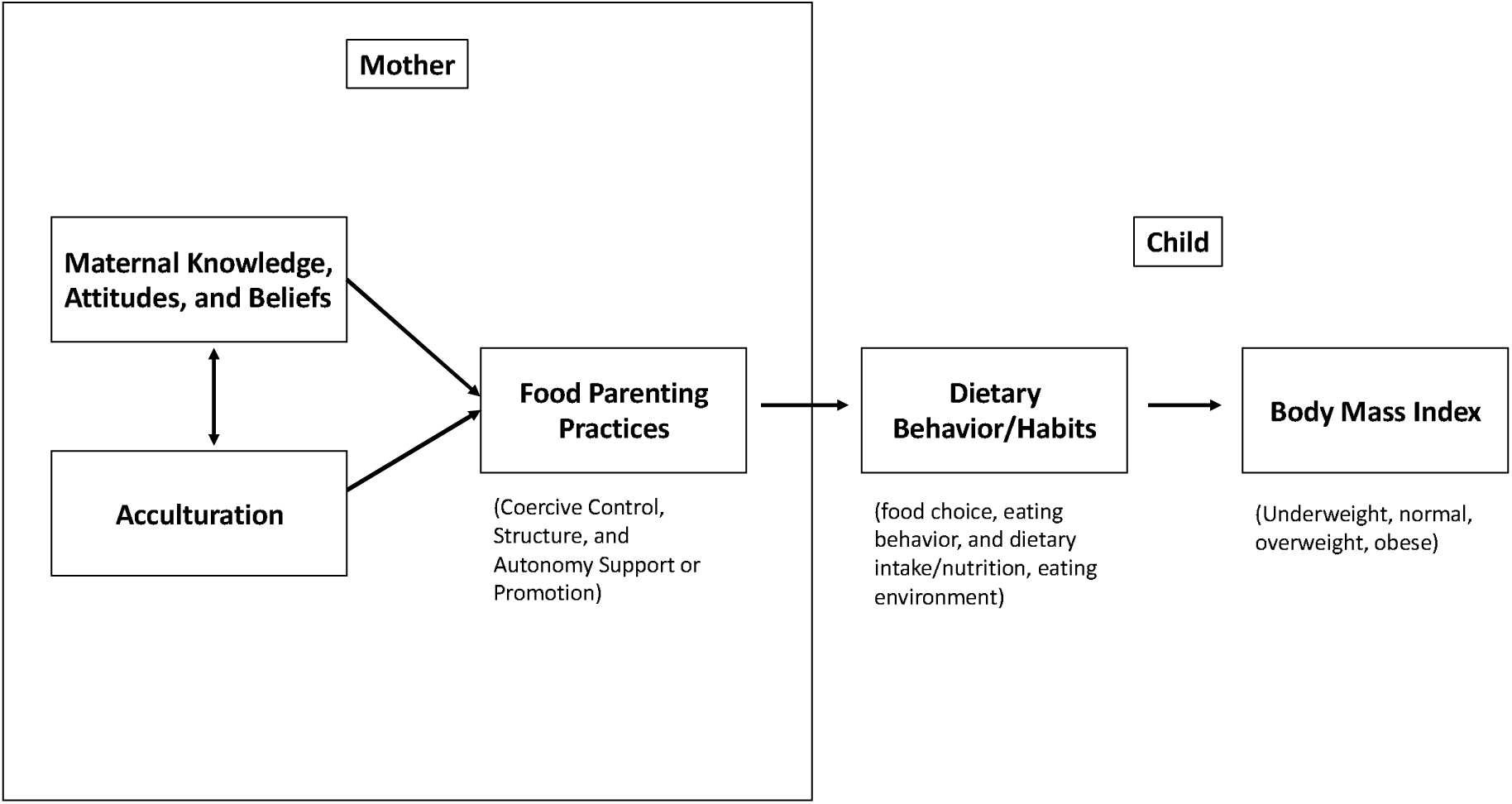
Proposed Model: Influences of Food Parenting Practices on BMI.

In turn, a mother’s food parenting practices affect the child’s dietary behavior and may impact BMI. SCT is particularly relevant for studying nutrition-related behaviors among Black immigrant mothers, whose personal (i.e., knowledge, attitudes, and beliefs) and environmental (i.e., acculturation) factors shape their behavioral (i.e., food parenting practices) practices, which in turn impact their children’s dietary behaviors.

Over the years, the number of people migrating to the U.S. has significantly increased. In particular, the Black immigrant population grew by 7% over 19 years, 2000-2019,^13^ making up 10% (4.6 million) of the U.S. population and projected to reach 9.5 million by 2060.^13^ This growing population includes children of Black immigrants who are often grouped in research alongside children of non-immigrant Blacks, despite potential distinct experiences and health risks. The increasing obesity rates among Blacks children and adolescents suggest an increased vulnerability for Black immigrant youth, and research focused on this subgroup remains limited. To date, few studies have explored the health behavior of Black immigrants in the U.S., and none have looked at the food parenting practices of Black immigrant mothers. Therefore, this study aims to examine the food parenting practices of Black immigrant mothers living in Metro Atlanta, GA using the Comprehensive Home Environment Survey (CHES), thereby addressing a critical gap in the literature and refining the proposed model, *Influences of Food Parenting Practices on BMI*, and offering a foundation for future quantitative research on child BMI.

## Methods

### Participant recruitment and selection

Black immigrant mothers were recruited via social media (i.e., Facebook, Reddit, Instagram, and WhatsApp) and personal networks between July 15, 2022, and August 23, 2022. A screening survey was used to determine study participants’ eligibility. Inclusion criteria were: female; older than 18 years old; born in an African, Caribbean, or Latin American country; fluent in English; Black, African, or Afro-Caribbean race or ethnicity; resides in one of the 11 metro Atlanta counties; has one or more children between the ages of 2 and 19; and is the primary caregiver of the child.

At the end of the survey, participants were directed to another screen to select a focus group to attend. Participants who completed the survey and attended a focus group received the study incentive, a $10 electronic gift card. Focus group findings are published elsewhere.^12^

### Measures

In this study, the Principal Investigator (PI) used the diet subscales of the Comprehensive Home Environment Survey (CHES) to measure the behavior of the child and mother. More specifically, the PI used the validated survey instrument Chen et al^14^ developed, a 40-item modified version of the original CHES, to assess food parenting practices and the comprehensive home environment. This survey includes scales related to three food parenting practices constructs (and their subconstructs): coercive control (restriction, pressure to eat, and threats and bribes), structure (meals and snack routines, modeling, rules and limits, and healthy food availability and accessibility), and autonomy support or promotion (child involvement).

### Data collection

Metro Atlanta, located in the southeastern US, is home to approximately 190,000 Black immigrants^13^. Data for this study were collected from Black immigrant mothers living in the metropolitan area of Atlanta, Georgia. The study was approved by the [BLINDED FOR REVIEW] on July 8, 2022.

After completing the screener survey, eligible participants were consented and invited to complete a brief survey with items assessing mother and child demographic data, country of birth, and length of residence in the US. After providing demographic data, participants were asked to answer a 44-question survey (40 CHES questions, one question comparing the youngest child’s feeding behavior to that of her other children, if applicable, and three questions from the 2003 New Immigrant Survey) with their youngest child in mind. Overall, the survey took 10-20 minutes to complete.

### Statistical Analysis

The PI conducted a univariate and bivariate descriptive analysis and linear regression. The univariate descriptive analysis provided the means with standard deviation (SD), median, mode, minimum, maximum, and frequencies, for the independent variables (e.g., sociodemographics) and the 40 items from the CHES survey. The bivariate analysis, independent t test, indicated the magnitude of association between the outcome variables (e.g., the food parenting practices subconstructs) and the six normally distributed independent variables (i.e., education, income, length of stay in the US, age of migration, child’s age, and child’s sex). The linear regression analysis predicted the value of the dependent variable based on the value of specific independent variables.

Prior to conducting the bivariate analysis to observe patterns and differences among study participants, the PI conducted a reliability test, to determine the level of internal consistency, and dichotomized the independent variables. First, to ensure all the CHES survey responses were on a continuous scale from 0-1, some items were reversed coded when necessary. Based on the reliability results, each subconstruct’s scale was adjusted (i.e., items were removed) where applicable, to obtain a Cronbach’s α of 0.70 or higher.^15^ Finally, the chosen six sociodemographics with a normal distribution – education (bachelor’s degree or less vs. graduate degree and higher), income (under $50,000 vs. $50,00 and above), years in the US (under 15 years vs. 15 years or more), age of migration (under age 14 vs. age 14 and older), child’s age (under age 5 vs. age 5 and older), and child’s sex (male vs. female) – were dichotomized to simplify the independent t test analysis and examine expected differences between the two distinct groups.^16^

Two subconstructs’ scales were excluded from the bivariate and linear regression analyses due to poor reliability: rules and limits (Cronbach’s α = 0.67) and restriction (Cronbach’s α = 0.69). Four subconstructs’ scales were modified to improve internal consistency. The meals and snack routines scale was modified to include four items instead of the original six, the modeling scale was shortened from seven items to four, the pressure to eat scale was shortened from four items to three, and the child involvement scale was modified to keep the planning items only. Table 1 represents the subconstructs’ scales and items included in the bivariate and explanatory analyses.

**Table 1.**
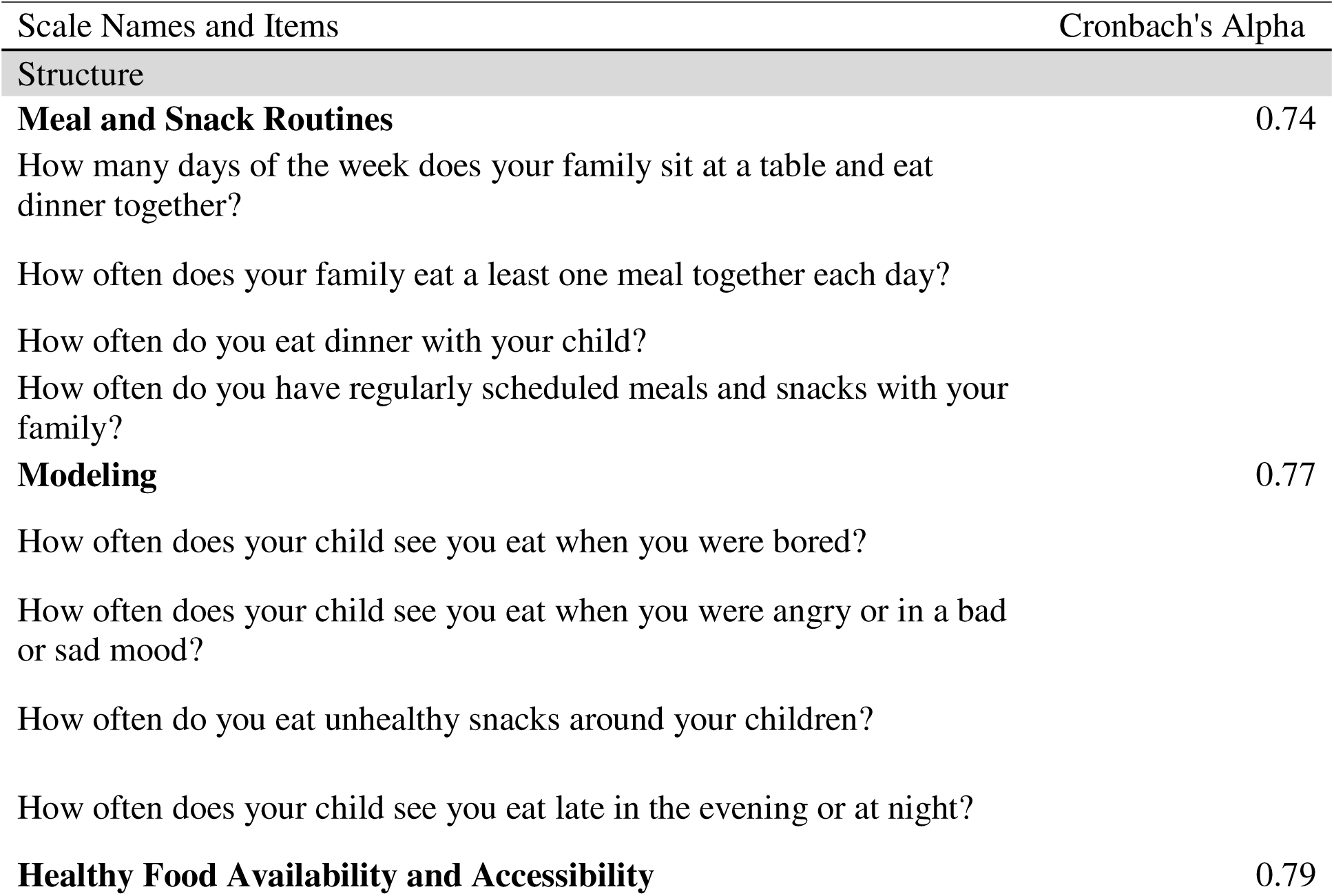
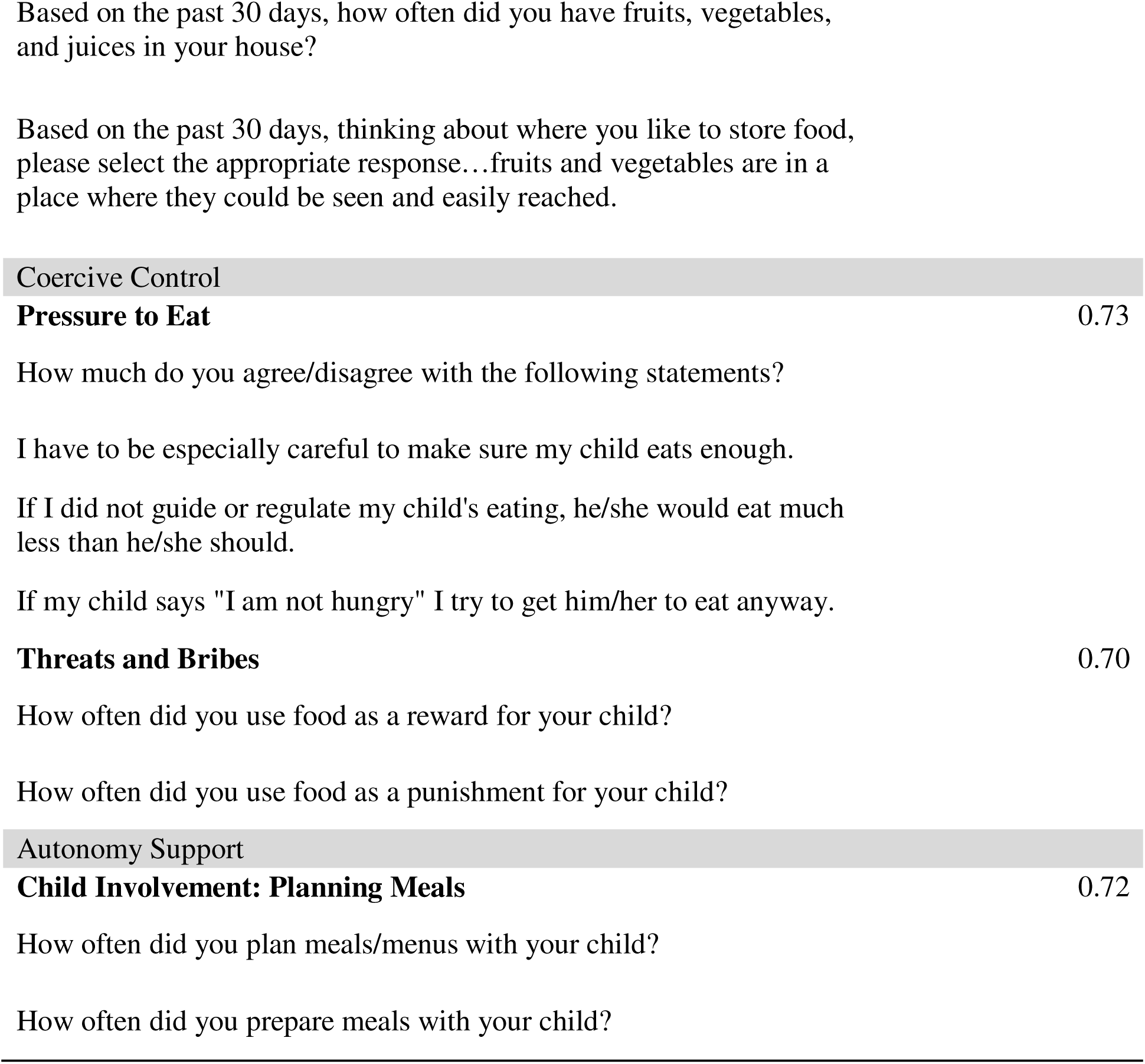
Food Parenting Practice Subconstructs Used in Bivariate and Explanatory Analyses.

After completing the reliability tests and adjusting the subconstructs’ scales accordingly, an independent samples t test was used to look for patterns and similarities between the six food parenting practice subconstructs’ scales and six sociodemographic variables. Variables that showed a statistically significant difference, where the p-value was .05 or less, were analyzed further in a simple linear regression to describe the relationships between the selected sociodemographic variables and corresponding food parenting practice subconstructs. Alpha was set at 0.05, therefore, p<0.05 was statistically significant. All quantitative statistical analyses were performed using the IBM SPSS Statistics (version 28.0.0.0).

## Results

The final analytic sample included 103 mothers who fit all inclusion criteria including, sex, age, country of birth, fluent in English, race/ethnicity, residing in Metro Atlanta, GA, with one or more children 2-19 years old, and being the primary caregiver of the child.

### Univariate descriptive analysis

#### Survey participants: sociodemographics

Most mothers were 25-34 years old (60.2%); non-Hispanic Black (93.2%), married (82.5%), worked outside the home (full-time 35.9%, part-time 30.1%), earned a household income of $100,000 per year (30.1%), had a bachelor’s degree (38.8%), and lived in the US for 15 years or more (48.5%). The average age of the mother’s youngest child was 5.6 ± 4.1 years, and 50.5% of children were male. Participants migrated to the US at an average age of 14.8 ± 10.1 years from Africa (77.7%), Latin America (10.7%), and the Caribbean (11.7%). See Table 2. Participants migrated from 32 countries, mostly Kenya (16.5%), Nigeria (20.4%), South Africa (10.7%).

**Table 2.** Survey Participants’ Sociodemographic Characteristics (n=103).

| Characteristics | n | % |
| --- | --- | --- |
| Mother |  |  |
| Age |  |  |
| 18-24 | 10 | 9.7 |
| 25-34 | 62 | 60.2 |
| 35-44 | 25 | 24.3 |
| 45-54 | 5 | 4.9 |
| prefer not to answer | 1 | 1 |
| **Race** |  |  |
| Non-Hispanic Black | 96 | 93.2 |
| Hispanic Black | 6 | 5.8 |
| prefer not to answer | 1 | 1 |
| **Education** |  |  |
| High school or less | 5 | 4.9 |
| Associate degree | 14 | 13.6 |
| Bachelor's degree | 40 | 38.8 |
| Graduate degree | 29 | 28.2 |
| Doctorate Degree | 15 | 14.6 |
| **Relationship** |  |  |
| Single and never married | 5 | 4.9 |
| Divorced | 1 | 1 |
| Separated | 5 | 4.9 |
| Widowed | 2 | 1.9 |
| Married | 85 | 82.5 |
| Single and living with a partner | 5 | 4.9 |
| **Occupation** |  |  |
| Full-time working outside the home | 38 | 36.9 |
| Part-time working outside the home | 30 | 29.1 |
| Working from home for a salary | 20 | 19.4 |
| Stay at home mom | 13 | 12.6 |
| prefer not to answer | 2 | 1.9 |
| **Income** |  |  |
| ≤\$49,999 | 42 | 40.8 |
| \$50,000 - <\$99,000 | 24 | 23.3 |
| >\$100,000 | 31 | 30.1 |
| prefer not to answer | 6 | 5.8 |
| **Children in Home** |  |  |
| 1 | 29 | 28.2 |
| 2 | 41 | 39.8 |
| 3 | 17 | 16.5 |
| 4 | 7 | 6.8 |
| 5 or more | 3 | 2.9 |
| Missing | 6 | 5.8 |
| **Region of Immigration** |  |  |
| Africa | 80 | 77.7 |
| Latin America | 11 | 10.7 |
| Caribbean | 12 | 11.7 |
| **Length in US** |  |  |
| < 5 years | 4 | 3.9 |
| 5- < 10 years | 22 | 21.4 |
| 10 - <15 years | 26 | 25.2 |
| 15 years or more | 51 | 49.5 |
| Child |  |  |
| **Youngest child's sex** |  |  |
| Male | 52 | 50.5 |
| Female | 51 | 49.5 |
| **Youngest child's age** |  |  |
| Under 5 | 57 | 55.3 |
| 5 and older | 46 | 44.7 |

#### Food parenting practices, weight concerns, and acculturation

Table 3 presents the results of the univariate descriptive analysis for the food parenting practices of Black immigrant mothers: coercive control (restriction, pressure to eat, and threats and bribes), structure (meals and snack routines, modeling, rules and limits, and healthy food availability and accessibility), and autonomy support or promotion (child involvement in planning meals and shopping).

**Table 3.**
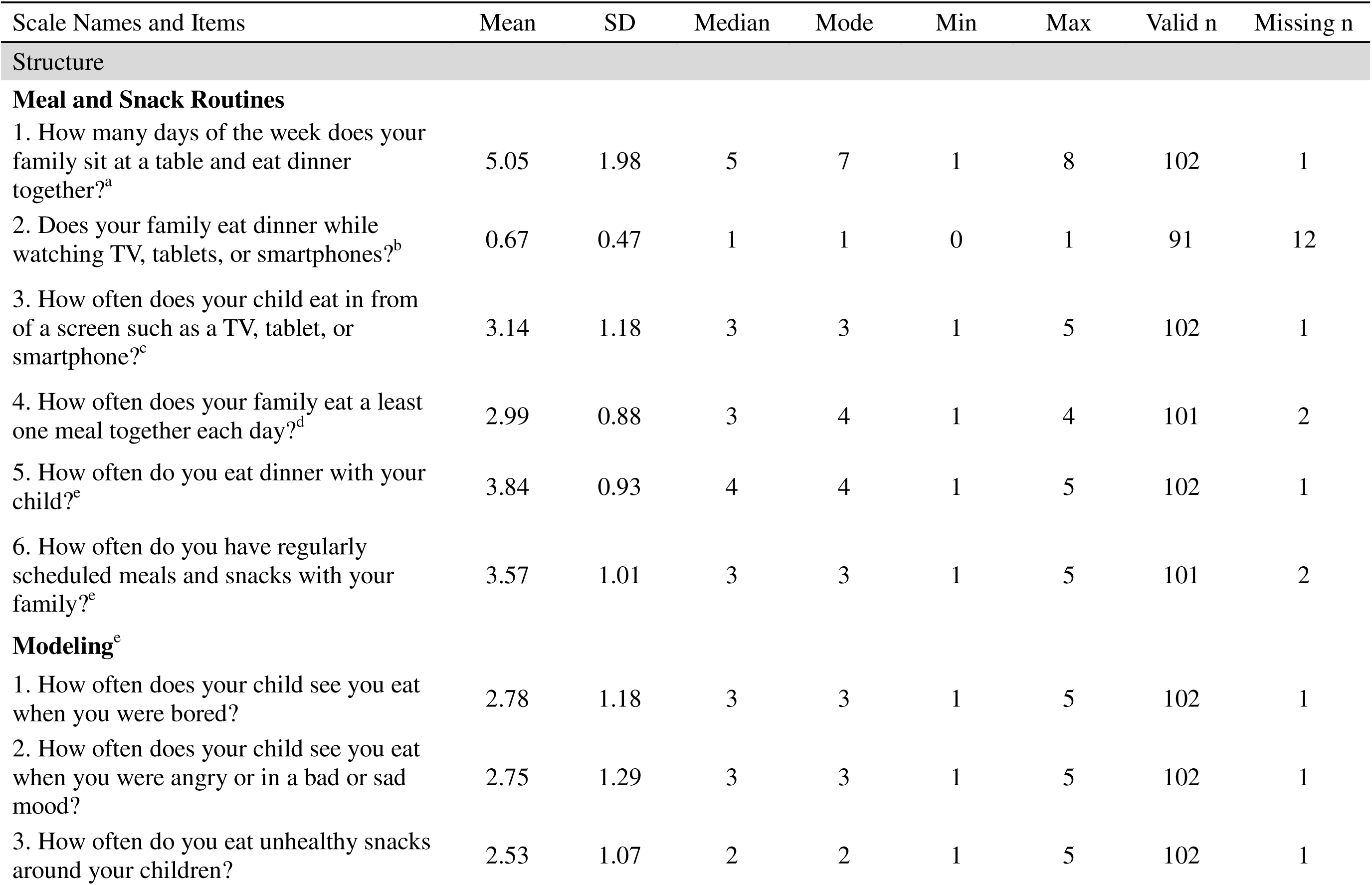
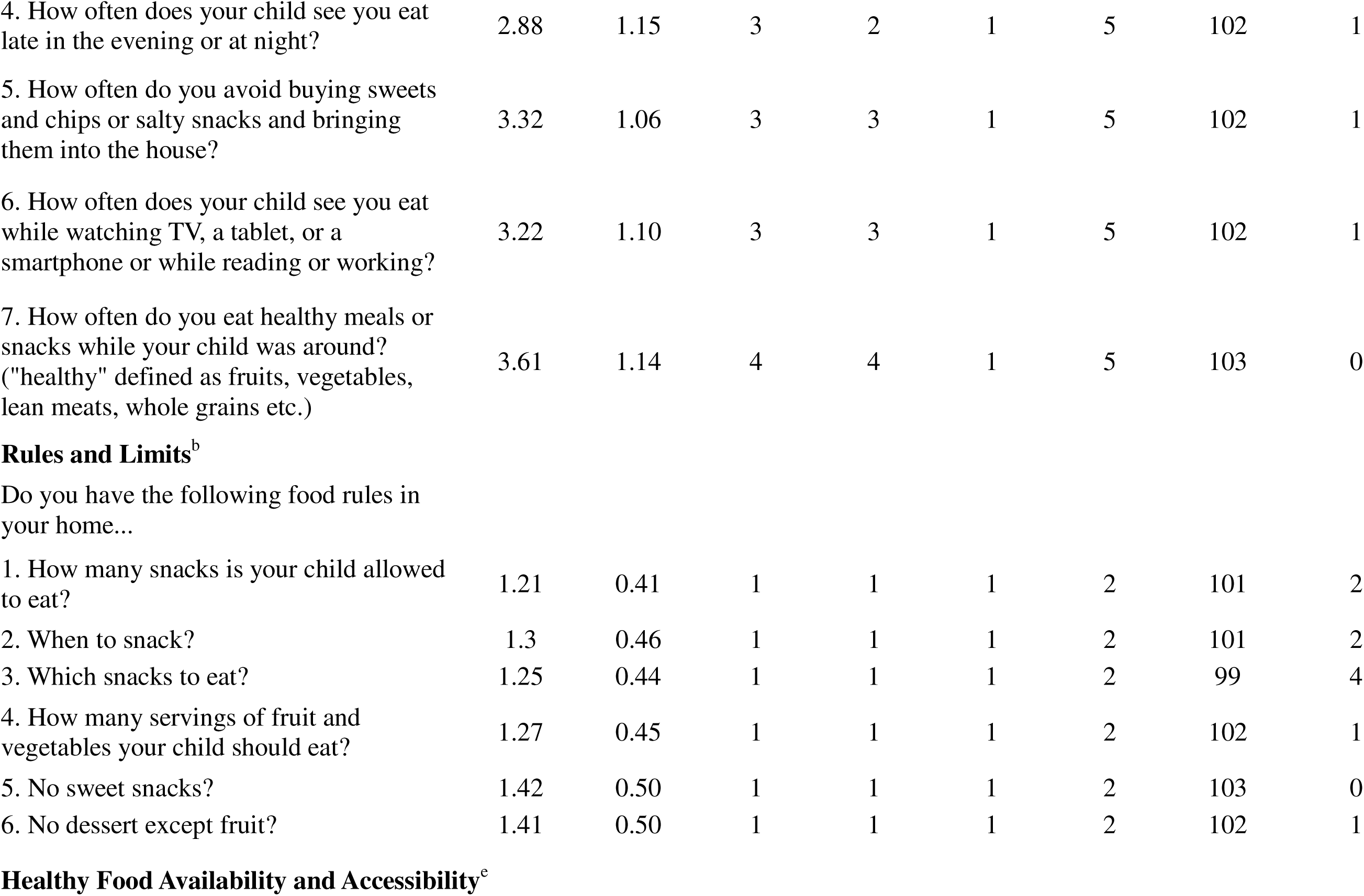
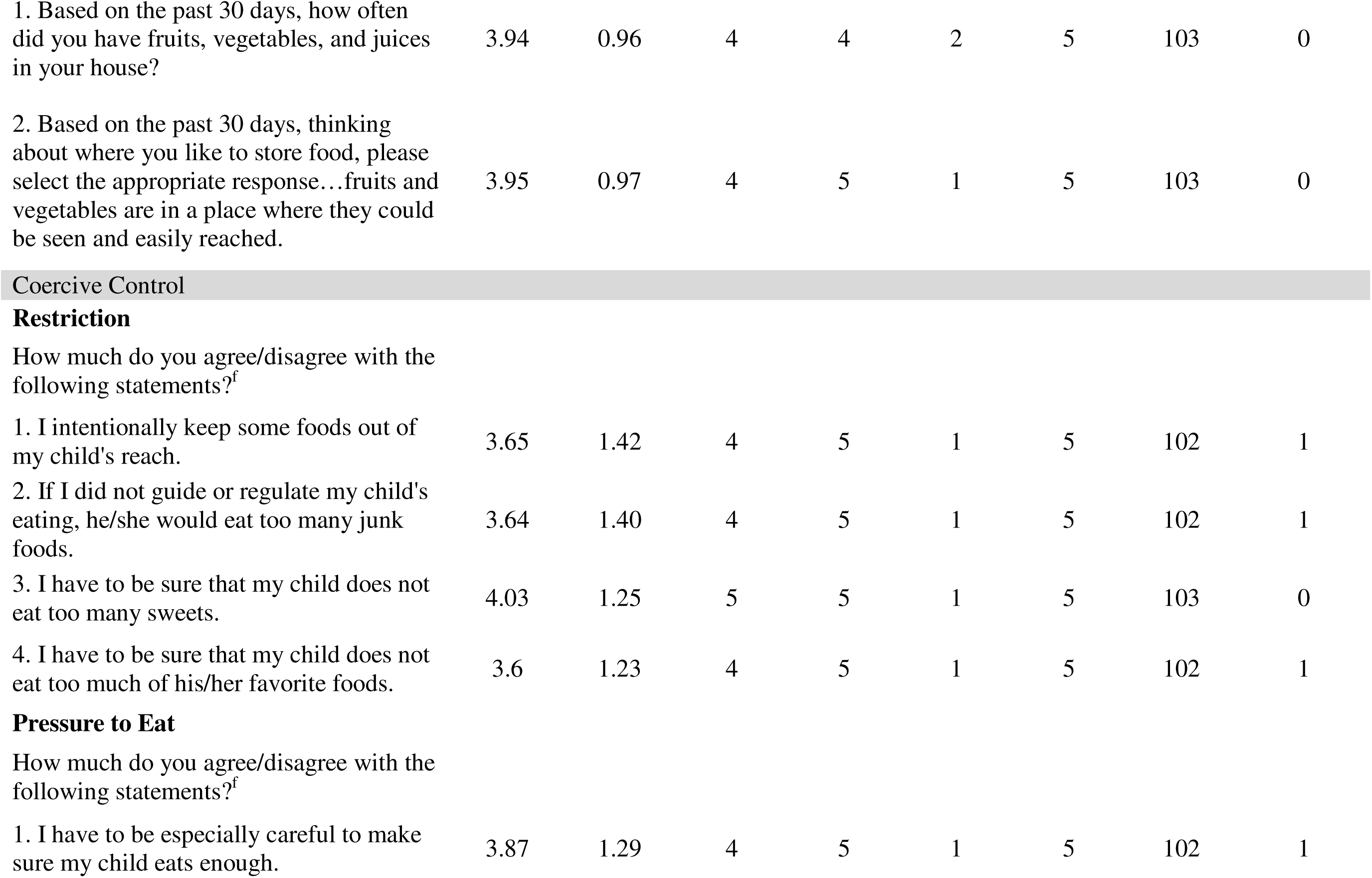
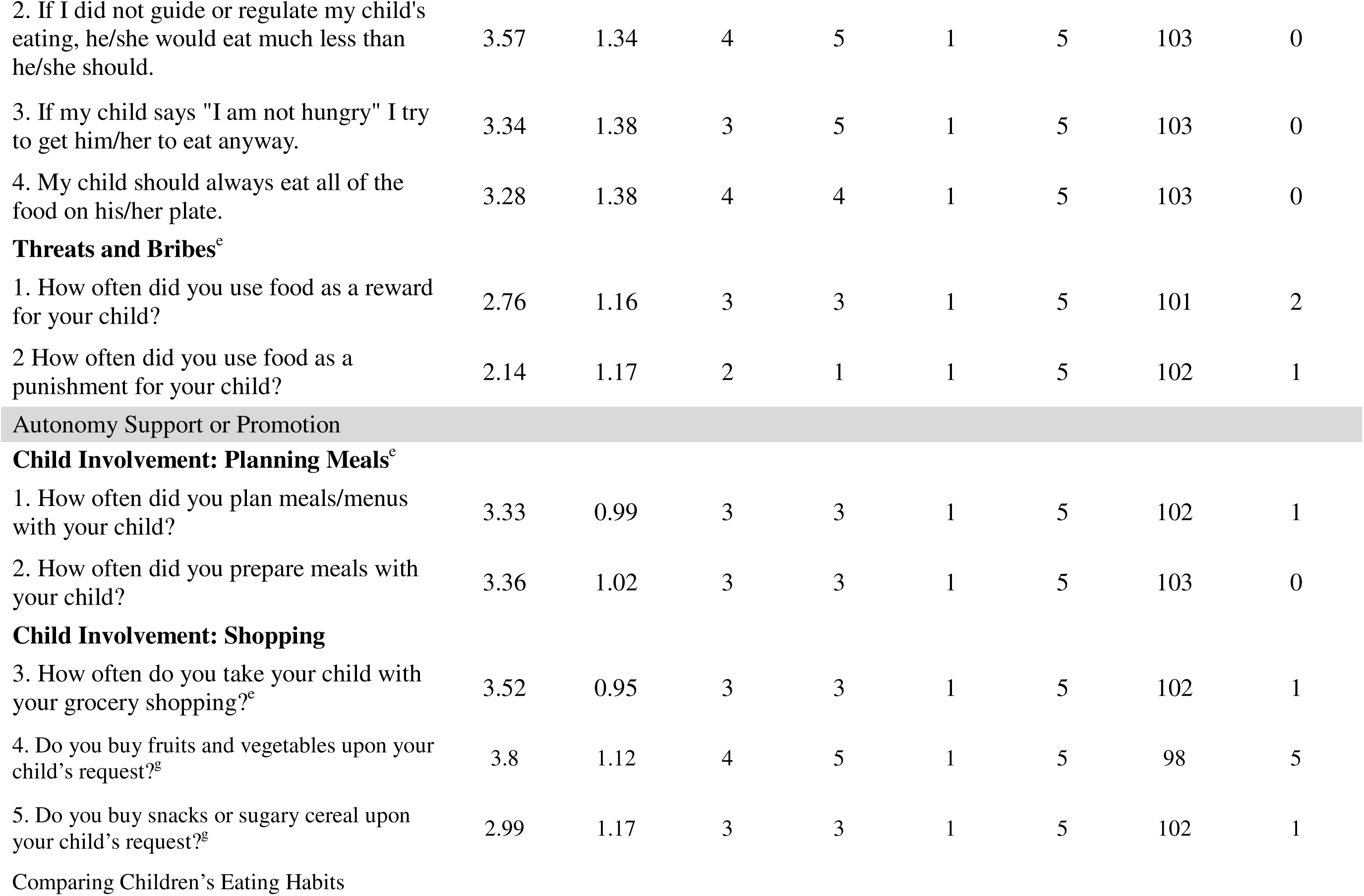
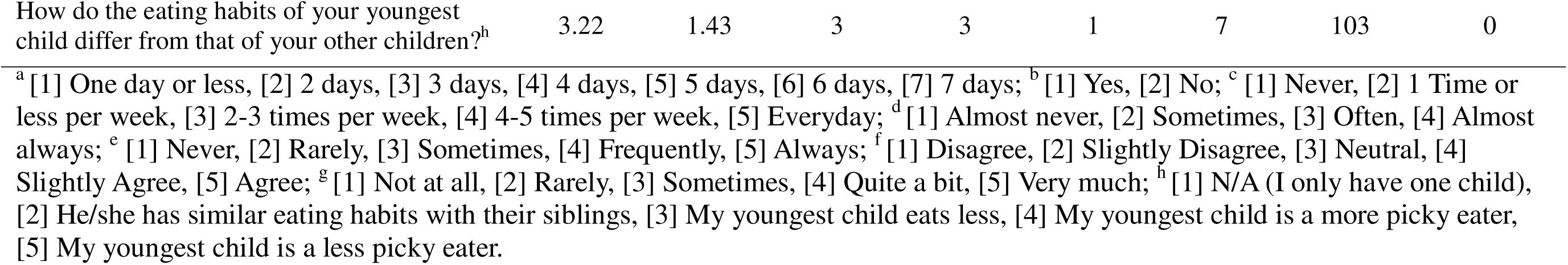
Univariate Analysis of CHES Scale Items.

*Coercive control*. Using a Likert-type agreement scale from 1 (disagree) to 5 (agree), participants indicated moderate agreement with items asking if they intentionally kept some foods out of the child’s reach, regulated junk food, and made sure the child does not eat too many sweets or their favorite foods. They also slightly agreed to pressuring their child to eat enough otherwise the child would eat much less and getting the child to eat even when he/she says, “I am not hungry”. Participants reported using food as a reward sometimes but less frequently as punishment.

*Structure*. Participants reported that their families ate dinner at a table together an average of 5 days out of the week, with 59.2% reporting watching tv or other devices while eating dinner. Mothers reported that their children saw them eat sometimes when bored, angry, sad, or in a bad mood, and eat unhealthy snacks. Participants indicated they ate healthy meals or snacks more frequently while their children were around. Concerning food rules in the home, 71.8% of participants had rules for the number of servings of fruits and vegetables the child should eat, 68.9% had rules regarding when to snack, and 71.8% had rules about which snack to eat. Over half of participants had rules prohibiting sweet snacks (58.3%) and serving fruits as dessert (58.3%). On a Likert-type scale from never (1) to always (5), mothers reported having healthy food available and accessible to their children frequently.

*Autonomy support or promotion*. Using a Likert-type frequency scale from 1 (never) to 5 (always), participants indicated that their children were sometimes involved in menu planning and meal preparation. Regarding shopping, mothers reported that they frequently took their child grocery shopping, frequently bought fruits or vegetables upon their child’s request, and sometimes bought snacks or sugary cereal when their child asked.

Because mothers answered the survey questions for only one child, their youngest, they were asked to describe how the eating habits of the youngest child differ from her other children to gain a more holistic view of her food parenting practices. For the 85% of participants with more than one child, 13.6% reported that the youngest child had similar eating habits to their siblings, 29.1% reported that the youngest child ate less, 23.3% reported they ate more, 12.6% reported that they youngest child was a pickier eater, and 4.9% reported they were a less picky eater.

*Weight concern.* Analysis of weight concern questions showed that generally, mothers were worried about their children’s weight. Using a Likert-type scale from 1 (unconcerned) to 5 (very concerned), participants indicated concern about their child having to diet to maintain a desirable weight; becoming overweight; eating too much when she is not around; and agreed to ‘I have to be sure that my child does not eat too much’. See Table 15.

*Acculturation*. When asked about their health before migrating to the US, 34% of mothers rated their health as “excellent” and 28.20% as “very good.” In comparison to their current health, less mothers rated their health as “excellent”, 27.20%, and more as “very good”, 36.90%. Overall, migrating to the US did not appear to have a negative impact on the health of these study participants, for 86.40% of mothers rated their current health as good or better. Interestingly, most mothers’ diets in the US were described as generally similar to what they ate in their native country. Using a scale of 1 (completely different) to 10 (exactly the same), participants indicated that their current diet was 6.15±2.24.

Overall, among the constructs assessed, there was greater use of pressure to eat, meal and snack routines, healthy food availability, and greater child autonomy. See Table 4.

**Table 4.**
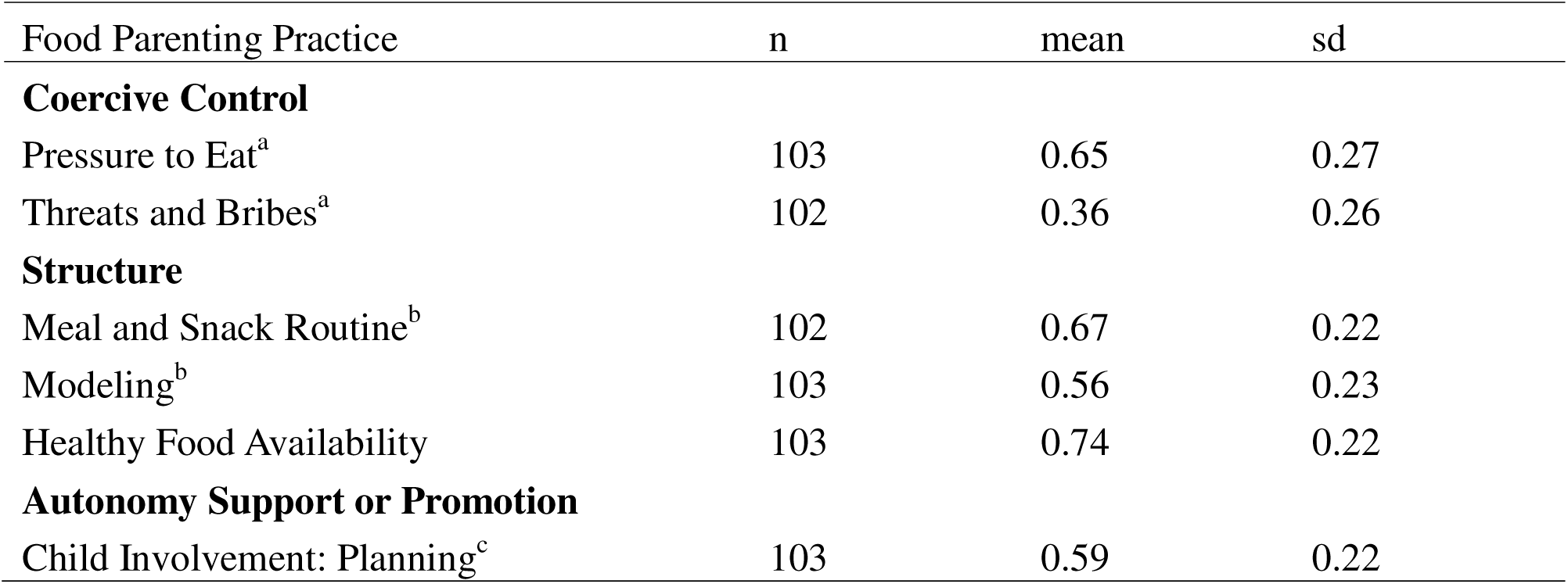
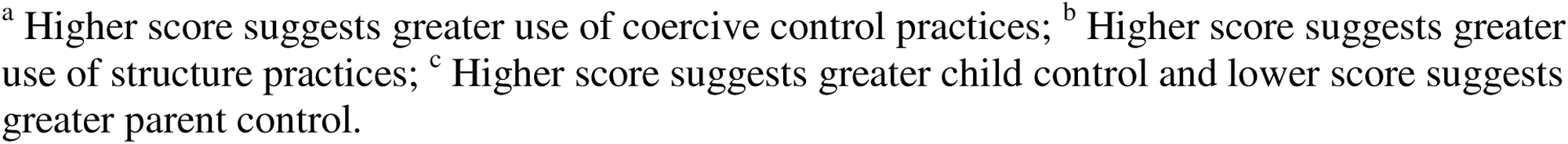
Food Parenting Practice Subconstruct Mean and Standard Deviation.

| Food Parenting Practice | n | mean | sd |
| --- | --- | --- | --- |
| **Coercive Control** |  |  |  |
| Pressure to Eat<sup>a</sup> | 103 | 0.65 | 0.27 |
| Threats and Bribes<sup>a</sup> | 102 | 0.36 | 0.26 |
| **Structure** |  |  |  |
| Meal and Snack Routine<sup>b</sup> | 102 | 0.67 | 0.22 |
| Modeling<sup>b</sup> | 103 | 0.56 | 0.23 |
| Healthy Food Availability | 103 | 0.74 | 0.22 |
| **Autonomy Support or Promotion** |  |  |  |
| Child Involvement: Planning<sup>c</sup> | 103 | 0.59 | 0.22 |

<sup>a</sup> Higher score suggests greater use of coercive control practices; <sup>b</sup> Higher score suggests greater use of structure practices; <sup>c</sup> Higher score suggests greater child control and lower score suggests greater parent control.

### Bivariate descriptive analysis – food parenting practices and weight

#### Coercive control

*Pressure to eat*. As presented in Table 5, an independent samples t-test showed no significant statistical differences between pressure to eat and all sociodemographic variables. However, the use of threats and bribes showed a statistically negative significant correlation for education (p = .042), income (p = .053, 95% confidence interval [-0.001, 0.200]), and years in the US (p = .026).

**Table 5.** Independent Samples Test – Assessing the Differences Between Sociodemographic and the Food Parenting Practice.

| Pressure to Eat |  |  |  |  |  | Threats and Bribes |  |  |  |  |
| --- | --- | --- | --- | --- | --- | --- | --- | --- | --- | --- |
| Sociodemographics | n | mean | sd | p | Cohens' d | n | mean | sd | p | Cohens' d |
| Education |  |  |  |  |  |  |  |  |  |  |
| ≤Bachelor | 59 | 0.61 | 0.23 | 0.12 | 0.27 | 58 | 0.41 | 0.23 | 0.042 | 0.25 |
| ≥Graduate degree | 44 | 0.7 | 0.3 |  |  | 44 | 0.3 | 0.28 |  |  |
| Income |  |  |  |  |  |  |  |  |  |  |
| <50K | 42 | 0.63 | 0.27 | 0.75 | 0.27 | 41 | 0.43 | 0.21 | 0.053 | 0.26 |
| ≥50K | 55 | 0.64 | 0.27 |  |  | 55 | 0.33 | 0.28 |  |  |
| Years in US |  |  |  |  |  |  |  |  |  |  |
| <15 years | 52 | 0.64 | 0.28 | 0.686 | 0.27 | 52 | 0.42 | 0.23 | 0.026 | 0.25 |
| ≥15 years | 51 | 0.66 | 0.26 |  |  | 50 | 0.3 | 0.27 |  |  |
| Age of Migration |  |  |  |  |  |  |  |  |  |  |
| <14 years old | 52 | 0.69 | 0.22 | 0.155 | 0.27 | 51 | 0.38 | 0.27 | 0.439 | 0.26 |
| ≥14 years old | 50 | 0.61 | 0.31 |  |  | 50 | 0.34 | 0.24 |  |  |
| Child's age |  |  |  |  |  |  |  |  |  |  |
| < 5 years | 57 | 0.68 | 0.25 | 0.177 | 0.27 | 57 | 0.33 | 0.28 | 0.199 | 0.26 |
| ≥5 years | 46 | 0.61 | 0.28 |  |  | 45 | 0.4 | 0.23 |  |  |
| Child's sex |  |  |  |  |  |  |  |  |  |  |
| Male | 52 | 0.64 | 0.26 | 0.778 | 0.27 | 52 | 0.38 | 0.25 | 0.495 | 0.26 |
| Female | 51 | 0.66 | 0.28 |  |  | 50 | 0.34 | 0.27 |  |  |

**Table 5.** *continued*
| Meal and Snack Routines |  |  |  |  |  | Modeling |  |  |  |  |
| --- | --- | --- | --- | --- | --- | --- | --- | --- | --- | --- |
| Sociodemographics | n | mean | sd | p | Cohens' d | n | mean | sd | p | Cohens' d |
| Education |  |  |  |  |  |  |  |  |  |  |
| ≤Bachelor | 58 | 0.65 | 0.22 | 0.23 | 0.223 | 59 | 0.54 | 0.21 | 0.164 | 0.23 |
| ≥Graduate degree | 44 | 0.7 | 0.22 |  |  | 44 | 0.6 | 0.25 |  |  |
| Income |  |  |  |  |  |  |  |  |  |  |
| <50K | 41 | 0.6 | 0.22 | 0 | 0.212 | 42 | 0.54 | 0.22 | 0.717 | 0.22 |
| ≥50K | 55 | 0.73 | 0.21 |  |  | 55 | 0.55 | 0.22 |  |  |
| Years in US |  |  |  |  |  |  |  |  |  |  |
| <15 years | 51 | 0.62 | 0.23 | 0.03 | 0.219 | 52 | 0.57 | 0.2 | 0.887 | 0.23 |
| ≥15 years | 51 | 0.72 | 0.2 |  |  | 51 | 0.56 | 0.26 |  |  |
| Age of Migration |  |  |  |  |  |  |  |  |  |  |
| <14 years old | 52 | 0.75 | 0.2 | <.001 | 0.21 | 52 | 0.5 | 0.23 | 0.009 | 0.22 |
| ≥14 years old | 49 | 0.59 | 0.22 |  |  | 50 | 0.62 | 0.21 |  |  |
| Child's age |  |  |  |  |  |  |  |  |  |  |
| < 5 years | 56 | 0.69 | 0.26 | 0.26 | 0.223 | 57 | 0.59 | 0.21 | 0.258 | 0.23 |
| ≥5 years | 46 | 0.64 | 0.17 |  |  | 46 | 0.54 | 0.24 |  |  |
| Child's sex |  |  |  |  |  |  |  |  |  |  |
| Male | 52 | 0.66 | 0.23 | 0.68 | 0.225 | 52 | 0.56 | 0.23 | 0.788 | 0.23 |
| Female | 50 | 0.68 | 0.22 |  |  | 51 | 0.57 | 0.23 |  |  |

**Table 5.** *continued*
| Healthy Food Availability and Accessibility |  |  |  |  |  | Child Involvement: Planning Meals |  |  |  |  |
| --- | --- | --- | --- | --- | --- | --- | --- | --- | --- | --- |
| Sociodemographics | n | mean | sd | p | Cohens' d | n | mean | sd | p | Cohens' d |
| Education |  |  |  |  |  |  |  |  |  |  |
| ≤Bachelor | 59 | 0.67 | 0.21 | <.001 | 0.21 | 59 | 0.56 | 0.2 | 0.095 | 0.88 |
| ≥Graduate degree | 44 | 0.83 | 0.19 |  |  | 44 | 0.62 | 0.25 |  |  |
| Income |  |  |  |  |  |  |  |  |  |  |
| <50K | 42 | 0.61 | 0.21 | <.001 | 0.2 | 42 | 0.54 | 0.2 | 0.082 | 0.88 |
| ≥50K | 55 | 0.81 | 0.18 |  |  | 55 | 0.62 | 0.23 |  |  |
| Years in US |  |  |  |  |  |  |  |  |  |  |
| <15 years | 52 | 0.69 | 0.23 | 0.05 | 0.22 | 52 | 0.56 | 0.19 | 0.328 | 0.89 |
| ≥15 years | 51 | 0.78 | 0.2 |  |  | 51 | 0.61 | 0.25 |  |  |
| Age of Migration |  |  |  |  |  |  |  |  |  |  |
| <14 years old | 52 | 0.73 | 0.2 | 0.79 | 0.22 | 52 | 0.61 | 0.24 | 0.388 | 0.89 |
| ≥14 years old | 50 | 0.74 | 0.24 |  |  | 50 | 0.57 | 0.21 |  |  |
| Child's age |  |  |  |  |  |  |  |  |  |  |
| < 5 years | 57 | 0.75 | 0.24 | 0.36 | 0.22 | 57 | 0.57 | 0.23 | 0.278 | 0.89 |
| ≥5 years | 46 | 0.71 | 0.19 |  |  | 46 | 0.6 | 0.21 |  |  |
| Child's sex |  |  |  |  |  |  |  |  |  |  |
| Male | 52 | 0.72 | 0.22 | 0.54 | 0.22 | 52 | 0.54 | 0.22 | 0.027 | 0.86 |
| Female | 51 | 0.75 | 0.22 |  |  | 51 | 0.63 | 0.22 |  |  |

| Weight Concerns |  |  |  |  |  |
| --- | --- | --- | --- | --- | --- |
| Sociodemographics | n | mean | sd | p | Cohens' d |
| Education |  |  |  |  |  |
| ≤Bachelor | 59 | 0.5 | 0.25 | 0.074 | 0.31 |
| ≥Graduate degree | 44 | 0.61 | 0.37 |  |  |
| Income |  |  |  |  |  |
| <50K | 42 | 0.62 | 0.24 | 0.113 | 0.3 |
| ≥50K | 55 | 0.53 | 0.33 |  |  |
| Years in US |  |  |  |  |  |
| <15 years | 52 | 0.56 | 0.28 | 0.707 | 0.31 |
| ≥15 years | 51 | 0.53 | 0.34 |  |  |
| Age of Migration |  |  |  |  |  |
| <14 years old | 52 | 0.63 | 0.29 | 0.008 | 0.3 |
| ≥14 years old | 50 | 0.47 | 0.31 |  |  |
| Child's age |  |  |  |  |  |
| < 5 years | 57 | 0.51 | 0.34 | 0.205 | 0.31 |
| ≥5 years | 46 | 0.59 | 0.27 |  |  |
| Child's sex |  |  |  |  |  |
| Male | 52 | 0.52 | 0.31 | 0.382 | 0.31 |
| Female | 51 | 0.57 | 0.32 |  |  |

*Threats and Bribes*. Overall, all mothers used threats and bribes rarely. Interestingly, mothers who had a graduate degree or higher, a yearly household income of $50,000 and above, or had lived in the US for 15 years or more had a statistically significant lower use of food as a reward or punishment with their children compared to mothers who had a bachelor’s degree or less, an annual household income below $50,000, or lived in the US for less than 15 years, respectively. See Table 5.

#### Structure

*Meals and Snack Routine*. It can be seen in Table 5, there was a statistically significant difference between mothers reporting high income and mothers reporting low income (p = .004), between mothers living in the US for less than 15 years and mothers living in the US for 15 years or more (p = .027), and between mothers who migrated to the US before age 14 and mothers who migrated to the US at age 14 or older (p < .001). Participants with an annual household income of $50,000 and above, participants who had lived in the US for 15+ years, and participants who migrated to the US before the age of 14 were more likely to use positive meal and snack routines (e.g., eating dinner as a family most weekdays, regularly scheduled meals and snacks) compared to those whose income was less than $50,000, those who had lived in the US for less than 15 years, and those who migrated to the US after the age of 14, respectively.

*Modeling*. Mothers who migrated to the US before the age of 14 modeled poor eating habits (e.g., eating when bored, sad, or angry, eating unhealthy snacks, or late at night) in front of their children less often than mothers who migrated at an older age (p = .009). See Table 5.

*Healthy Food Availability and Accessibility*. Participants with a graduate degree or higher, participants with an annual household income of $50,000 or more, and participants who had lived in the US for 15 years or more were more likely to have healthy food available and accessible for their children (education p < .001, income p < .001, years in the US p = .049) compared to those whose income was below $50,000, and those who had lived in the US for less than 15 years. See Table 5.

#### Autonomy Support or Promotion

*Child Involvement: Planning Meals*. As seen in Table 5, there was one significant relationship in child involvement; female children were more involved in planning meals (p = .03).

#### Weight Concern

In a bivariate analysis, weight concern showed a statistically significant difference for age of migration (*p* = .008). Mothers who migrated before the age of 14 were more concerned about their child’s weight than mothers who migrated at age 14 and older.

### Explanatory Analysis – Simple and Multiple Linear Regression

A one-way ANOVA revealed that four sociodemographic variables influenced three food parenting subconstructs. Specifically, income, age of migration, and years in the US, could be used to reliably predict meals and snack routine (F (3, 92) = [8.22], p < .001). Education, income, and years in the US, could be used to reliably predict healthy food availability and accessibility (F (3, 93) = [13.51], p < .001), and threats and bribes (F (3, 92) = [2.84], p = .042). See Table 6.

**Table 6.** Simple and multiple linear regression for threats and bribes, meals and snack routines, modeling, and healthy food availability and accessibility.

| **Coercive control: Threats and Bribes** |  |  |  |
| --- | --- | --- | --- |
| Variables | B | t | p |
| (Constant) | 0.69 | 6.05 | <.001 |
| Education | -0.08 | -1.4 | 0.17 |
| Income | -0.06 | -1.12 | 0.27 |
| Years in US | -0.08 | -1.45 | 0.15 |
| **Structure: Meal and Snack Routines** |  |  |  |
| Variables | B | t | p |
| (Constant) | 0.7 | 6.01 | <.001 |
| Income | 0.12 | 2.72 | 0.01 |
| Years in US | 0.01 | 0.22 | 0.83 |
| Age of migration | -0.15 | -3.53 | <.001 |
| **Structure: Modeling** |  |  |  |
| Variables | B | t | p |
| (Constant) | 0.39 | 5.64 | <.001 |
| Age of Migration | 0.12 | 2.67 | 0.01 |
| **Structure: Healthy Food Availability and Accessibility** |  |  |  |
| Variables | B | t | p |
| (Constant) | 0.23 | 2.75 | 0.01 |
| Education | 0.14 | 3.6 | <.001 |
| Income | 0.16 | 4.03 | <.001 |
| Years in the US | 0.03 | 0.64 | 0.53 |
| **Autonomy Support or Promotion: Child Involvement: Planning Meals** |  |  |  |
| Variables | B | t | p |
| (Constant) | 0.44 | 6.55 | <.001 |
| Child's Sex | 0.1 | 2.25 | 0.03 |

Threats and bribes. R Square = 0 .085; Meals and snack routine. R Square = 0.211; Healthy food availability and accessibility. R Square = 0.303.

#### Coercive Control

*Threats and Bribes*. A multiple linear regression was run to predict threats and bribes used by mothers, from education, income, and years in the US. The model did not statistically significantly predict the use of threats and bribes. In addition, there was no evidence of multicollinearity among the variables that comprised the model, as the variance inflation factor (VIF) for each variable was between 1.06 and 1.14. See Table 6.

#### Structure

*Meals and Snack Routine*. In a multiple linear regression model examining relationships with meals and snack routines score, income was significantly related to meals and snack routines (p = .01) when adjusting for age of migration and length of stay in the US. See Table 6.

Individuals with an annual family income of $50,000 and above were more likely to use meals and snack routines (higher scores reflect greater use of this subconstruct/practice) compared to those with an annual family income below $50,000 (adjusted mean score 0.71 and 0.59, respectively). Also, age of migration was statistically significantly related to meals and snack routines (p < .001) when adjusting for income and length of stay in the US. Mothers who migrated before the age of 14 were more likely to use positive meals and snack routines compared to those who migrated at the age of 14 or older (adjusted mean score 0.73 and 0.58 respectively). Also, a multicollinearity problem did not exist in the model as VIF for each variable was between 1.10 and 1.25. See Table 6

*Modeling*. A simple linear regression model to explore the relationship between the dependent variable modeling and independent variable age of migration showed a statistically significant relationship. The age of migration statistically significantly predicted the mother’s modeling behavior (p = .01). Mothers who migrated to the US at the age of 14 or older modeled less healthy eating habits compared to mothers who migrated before age 14 (adjusted mean 0.62 and 0.50, respectively). See Table 6.

*Healthy Food Availability and Accessibility*. In a multiple linear regression model examining the relationship with healthy food availability and accessibility score, mother’s education was statistically significantly related to healthy food availability and accessibility (p < .001) when adjusting for income and years in the US. Mothers with a graduate degree or higher showed a greater use of this food parenting practice subconstruct compared to mothers with a bachelor’s degree or lower (adjusted mean 0.78 and 0.65, respectively). Additionally, income was also statistically significantly related to healthy food availability and accessibility (p < .001) when adjusting for education and years in the US. Households with an annual income of $50,000 and above had healthy food available and accessible in their homes more frequently compared to households with an income below $50,000 a year (adjusted mean 0.81 and 0.63, respectively). Furthermore, a multicollinearity problem did not exist in the model as VIF for each variable was between 1.06 and 1.13. See Table 6.

#### Autonomy Support or Promotion

*Child Involvement: Planning Meals*. In a simple linear regression model examining the relationship with child involvement in planning meals, child’s sex was significantly related to the dependent variable (p = .03). Female children were more likely to be involved in planning meals compared to male children (adjusted mean score 0.64 and 0.54, respectively). See Table 6.

#### Weight Concern

A simple linear regression model examining the relationship with weight concern score, confirmed that age of migration statistically significantly predicted the mother’s weight concern (*p* = .001). Individuals who migrated before the age of 14 are more concerned about their child having to diet to maintain a desirable weight, becoming overweight, and eating too much when she is not around compared to those who migrated at age 14 or older (adjusted mean 0.63 and 0.47 respectively).

#### Acculturation

When asked about their health before migrating to the US, 34% of mothers rated their health as “excellent” and 28.20% as “very good.” In comparison to their current health, less mothers rated their health as “excellent”, 27.20%, and more as “very good”, 36.90%. Overall, migrating to the US did not appear to have a negative impact on the health of these study participants, for 86.40% of mothers rated their current health as good or better. Interestingly, most mothers’ diets in the US were described as generally similar to what they ate in their native country. Using a scale of 1 (completely different) to 10 (exactly the same), participants indicated that their current diet was 6.15±2.24.

## Discussion

The purpose of this study was to learn more about the food parenting practices among a group of Black immigrant mothers who reside in Metro Atlanta, GA and to inform the *Influences of Food Parenting Practices on BMI* model based on survey data and previously published focus group transcripts.^12^ The present research was designed to fill an existing gap in the literature about the food parenting practices of this specific population in the US, and its influence on their children’ dietary behaviors. This study was guided by the social cognitive theory and the Influences of Food Parenting Practices on BMI model, and it provided insight into the knowledge, attitudes, and beliefs of Black immigrant mothers on nutrition. Additionally, the study clarified the types of food parenting practices some Black immigrant mothers use, the dietary behaviors and habits of their children, and acculturation’s role on their nutrition overall.

The results of this study indicate that Black immigrant mothers use all three food parenting practice constructs, coercive control (e.g., restriction, pressure to eat, threats and bribes), structure (e.g., rules and limits, meals and snack routines, modeling, food availability and accessibility), and autonomy support or promotion (e.g., child involvement).

### Coercive Control

Under coercive control, there is a good use of restriction among Black immigrant mothers. This finding matches what ^17^ observed in their cross-sectional study of 3 different cultures, where Black Afro-Caribbean families in the United Kingdom used restriction the most. Also, Power et al.^18^ found that first generation Hispanic immigrant mothers were more likely to use restriction with their young children. In focus group discussions by Tchoua et al.^12^, some mothers eliminate sugar from their child’s diet, give them fruit instead of junk food, and are selective on which processed food the child can eat. Their education, knowledge, and experience on nutrition inform those decisions; mothers attribute tooth decay to candy, describe junk food as unhealthy, and homecooked meals as the best option overall^12^.

Pressure to eat is another coercive control food parenting practice used among this study population. On average, mothers pressured their children to eat enough, even when “not hungry”, and to “clean their plate.” These results confirm what mothers shared during the focus group discussions in Tchoua et al.^12^ Because mothers believe that a child’s diet is instrumental in his or her growth and development, they insist on the child eating what she provides. In contrast to earlier findings by Kengneson et al,^19^ there was no significant correlation between different groups of mothers when divided by income, and years in the US, and use of pressure to eat. Both groups showed similar average scores for pressure to eat.

Threats and bribes are the least used food parenting practice. In the present investigation, preliminary analyses suggested that mothers who have a bachelor’s degree or less, a household income under $50,000 a year, lived in the US for less than 15 years, reported a greater use of threats and bribes with their children compared to other mothers. These preliminary findings were not confirmed in a multiple linear regression model.

### Structure

The survey results of this study indicate that Black immigrant families sit at a table and eat dinner as a family about five days a week, often have at least one meal daily together, sometimes have regularly scheduled meals and snacks as a family, and mothers frequently eat dinner with their children. This finding also corresponds with focus group findings where mothers noted having a less structured eating schedule since moving to the US^12^. In their native countries, eating three square meals a day was described as a common practice.

Concerning rules and limits, over 50% of mothers responding to the survey have food rules in their home for various food items (i.e., number of fruits and vegetables to consume, no sweet snacks, fruit as dessert, and the what/when/how many of snacks). This finding can be clarified by the mothers’ knowledge, attitudes, and beliefs shared during the focus group interviews^12^. Overall, mothers want their children to eat less sugar, junk, and processed foods because they believe these foods have a negative impact on health. Therefore, it is not surprising that healthy food was frequently available and accessible in the home, especially those with more resources.

Mothers with more education, household income, and years in the US were significantly more likely to use available and accessible healthy food compared to mothers with lower education level, household income, and years spent in the US. A multiple linear regression showed that education and income did positively influence healthy food availability and accessibility. This finding aligns with previous research that showed that financial resources impacted food choices. Immigrants with less financial resources made poor food choices^20–23^ and tended to have more access to unhealthy foods and less access to healthy food, because cost is a barrier.^24^

Surprisingly, despite talking consistently about the importance of healthy eating, mothers frequently modeled unhealthy eating behaviors, such as eating when bored, angry, sad, or in a bad mood, eating unhealthy snacks around their children, or eating late at night. However, in focus group interviews, several mothers mentioned eating more nutrient-dense foods (i.e., whole foods, unprocessed or minimally processed food) in front of their children, and having unhealthy foods away from them because they wanted the child to have better eating habits. Some avoid buying ‘super processed’ foods, while only one mother did not avoid any food.^12^

### Autonomy Support or Promotion

The survey results indicated that mothers sometimes planned and prepared meals/menus with their children. When grocery shopping, they sometimes take the child with them and are more likely to buy fruits and vegetables upon the child’s request versus snacks or sugary cereal. These findings can be further illuminated by the focus group themes of knowledge, attitudes, beliefs, modeling, and child’s diet. Most mothers want to reduce their child’s consumption of sugar and snacks. When asked to share about their child’s diet, fruits and vegetables were staple items and more common than sugary cereal.^12^

While the final construct of the proposed model (relationship to child BMI) could not be tested, the qualitative findings presented in this work offer promise for future quantitative investigation of this model and the SCT-derived elements that may predict child BMI. Mother’s Concern for Child’s Weight. This study demonstrates a correlation between the mother’s age of migration and her concern for the child’s weight. According to survey findings, mothers who migrated to the US before age 14 were more concerned about their child’s weight in comparison to mothers who relocated to the US at age 14 or older. This concern is warranted as previous research has shown that rates of overweight and obesity in the US have continually increased since the 1970s and Black children have a higher prevalence of being overweight or obese compared to White children. Immigrants who migrate to the US at a younger age are more susceptible to obesity as their duration in the US increases.^25^ The level of acculturation of a parent can impact the BMI of their child. As Latino mothers’ stay in the US increased, they adopted the American Culture more, and their children’s BMI increased overtime.^26^ Previous studies reported an increase in obesity as stay in the US increases and between diet acculturation and diet-related disease in sub-Saharan Africans living in the US.^21,27,28^

### The Influences of Food Parenting Practices on BMI Model

Following the analysis of survey responses and previously published focus group transcripts^12^, the proposed *Influences of Food Parenting Practices on BMI* model was revised accordingly. Most hypothesized relationships were supported, with several new pathways emerging and one, linking dietary habits to child BMI, remaining unvalidated due to unreliable self-reported anthropometric data. (See Fig 2)

**Fig 2.**
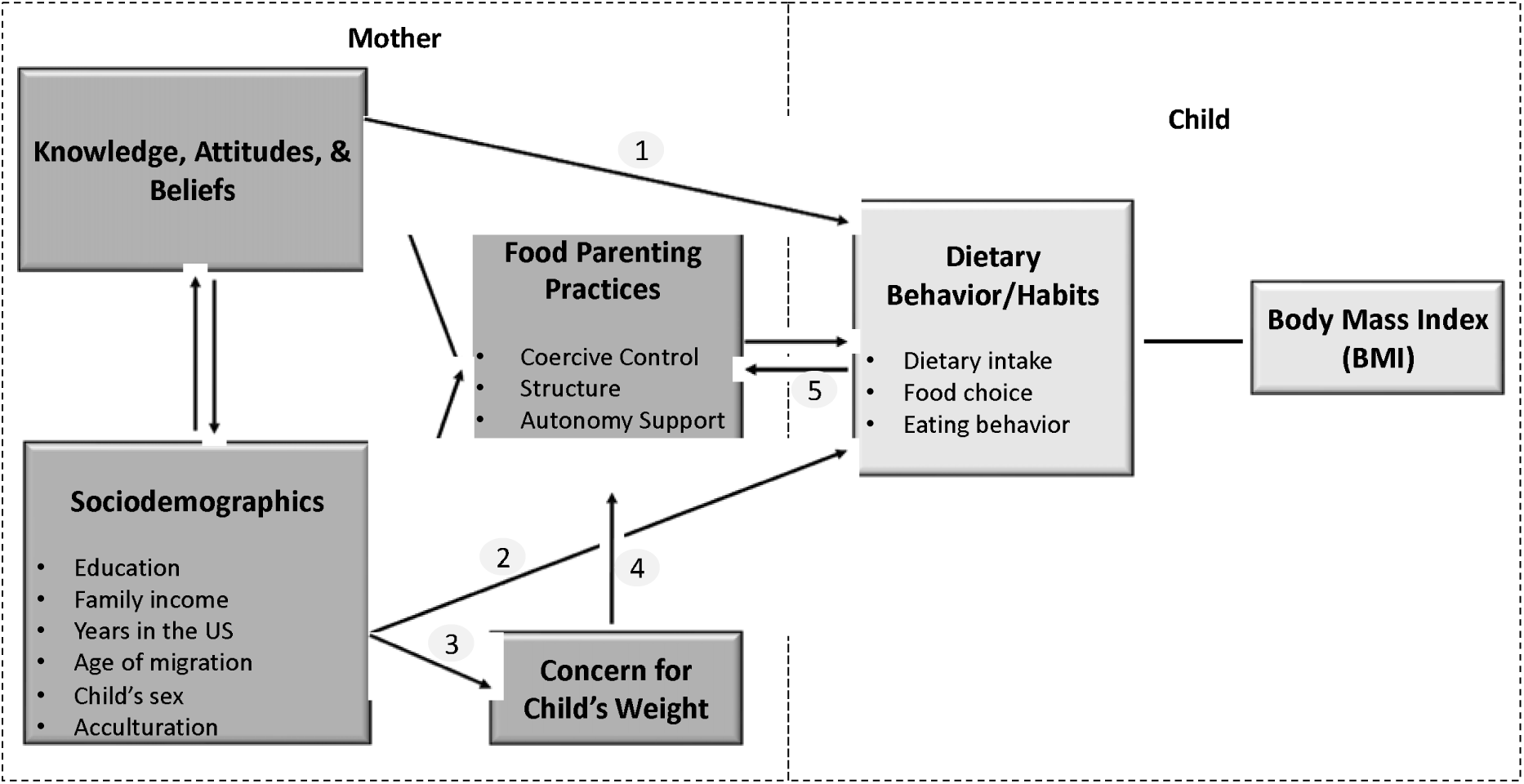
Influences of Food Parenting Practices on BMI Model. Conceptual model illustrating the influences on children’s dietary habits and maternal food parenting practices. Children’s dietary behaviors are shaped by maternal food parenting practices, as well as maternal knowledge, attitudes, and beliefs (KABs) (arrow 1), and sociodemographic factors (arrow 2). Maternal concern for the child’s weight influences her food parenting practices (arrow 4) and is itself shaped by sociodemographic factors (arrow 3). A reciprocal relationship exists between the child’s dietary habits and maternal food parenting practices (arrow 5). Arrows 1 is based on focus group findings^12^; arrow 4 is supported by survey data; and arrows 2, 3, and 5 are informed by both survey and focus group data.

#### Mother’s Knowledge, Attitudes, and Beliefs (KABs)

The current study confirms that a reciprocal relationship between KABs and sociodemographics exists and the KABs influences mothers’ food parenting practices. Analysis of focus group^12^ data found that the maternal KABs was also related to parental perceptions of the child’s dietary behavior or habit.

#### Mother’s Sociodemographics

Survey and focus group^12^ findings revealed that the mother’s education, family income, years in the US, age of migration, child’s sex, and acculturation affect her food parenting practices, her concern for child’s weight, and the child’s dietary behavior and habits.

#### Mother’s Food Parenting Practices

A central focus of this study was to learn more about the food parenting practices of Black immigrant mothers living in metro Atlanta, GA. Survey results of this study showed that Black immigrant mothers tended to use *coercive control* (i.e., restriction, pressure to eat, threats and brides), *structure* (i.e., meals and snack routines, modeling, rules and limits, healthy food availability and accessibility), and *autonomy support or promotion* (i.e., child involvement) in feeding their children, which directly influences their children’s dietary behavior and habits. Specifically, these mothers use of rules and limits, healthy food availability and accessibility, and restriction of unhealthy foods were found to influence the child’s food choice. Pressure to eat, meals and snack routines, modeling, and child involvement directly affect dietary intake. In fact, there is a reciprocal relationship between the mother’s food parenting practices and the child’s dietary behavior. Mother’s perceptions of their children’s dietary behavior and habits influence the food parenting practice the mother uses. Focus group discussions confirmed the mother’s use of coercive control and structure to influence the child’s dietary habits.^12^

#### Mother’s Concern for Child’s Weight

An interesting finding from survey data is the influence of the mother’s sociodemographics on her concern for her child’s weight, and that concern affects her food parenting practices.

#### Child’s Dietary Behavior/Habits

The present study and focus group^12^ findings identified two main maternal influences on the child’s dietary behavior or habits, mother’s KABs and sociodemographics.

The findings in this report are subject to a few limitations. First, 103 participants met all study inclusion criteria. Due to a small sample size, these findings cannot be extrapolated to all Black immigrants living in the US. Second, participants’ SES level was generally noted to be moderately high. A significant portion of this study’s population had a high-income level (i.e., 53% earned $50,000 or more) and a high educational level (i.e., 81.6% had a bachelor’s degree or higher). These percentages, of mothers in metro Atlanta, Georgia, might not be representative of average yearly income and education level of all Black immigrant mothers in other geographic areas in the US. Finally, social desirability bias, is a frequently noted limitation found in focus groups and self-reported survey data.^29^ Likewise, this study is not exempt from this bias, which should be considered when interpreting the study’s findings. Overall, there is a great need for more studies among Black immigrants in the US, especially among those who migrated to the U.S. voluntarily and are part of a lower SES status. Notably, more food parenting practice data is needed for Black immigrants from the wide range of sociodemographic background (i.e., from groups of low, middle, and high income and educational levels). Notwithstanding these limitations, this study addressed an existing gap in the literature on the food parenting practices used by Black immigrant mothers and serves as a base for future studies.

## Data Availability

All data produced in the present study are available upon reasonable request to the authors.

